# Heterogeneity in prognostic value of the neutrophil-to-lymphocyte ratio: a meta-analysis

**DOI:** 10.1101/19011387

**Authors:** Rachel Howard, Peter A. Kanetsky, Kathleen M. Egan

## Abstract

High pre-treatment values of the neutrophil-to-lymphocyte ratio (NLR) are strongly associated with poorer survival outcomes in cancer patients. Here, we assess heterogeneity in the magnitude of this association and the prognostic potential of the NLR between patient subgroups. We conducted a random effects meta-analysis of 228 published studies (N=75,555 patients) relating NLR with overall survival across 18 cancer types. Cochran’s Q test and Higgins I2 statistic were used to assess study heterogeneity. Pooled hazard ratios were compared between groups of studies classified by cancer type, geographic region, therapy type, and cut-off for high NLR to identify study-level characteristics associated with increased prognostic potential of the NLR. Pooled hazard ratios are highest in studies of melanoma and breast cancer and lowest in studies of brain cancer and lung cancer. Radiation as primary treatment also demonstrates a large pooled effect size as compared to other therapies. The NLR has greater prognostic value in certain cancer types and therapeutic regimens. Efforts are needed to comprehensively examine populations in which NLR has maximum prognostic power. Clinically meaningful thresholds for risk stratification should be identified within these patient subgroups to permit prospective validation of the prognostic potential of the NLR.

## 1. Introduction

Inflammation plays a key role in the pathophysiology of cancer^1-4^, and there is a strong interest in identifying easily obtainable and robust metrics of the systemic inflammatory status of cancer patients for risk stratification at diagnosis. Immune cell components of the complete blood count (CBC) offer an easily accessible measure of inflammation as the CBC is often collected as part of standard clinical care at minimal cost and inconvenience to the patient.

One marker of systemic inflammation available from the CBC is the neutrophil to lymphocyte ratio (NLR), the quotient of the absolute neutrophil and lymphocyte counts^5-8^. Neutrophilia is a common feature of cancer-associated chronic inflammation, and both tumor-promoting and immune-suppressive roles of neutrophil subpopulations have been documented^9-13^. In addition to producing cytokines associated with tumor progression, the innate neutrophilic response can suppress the activity of cytotoxic T cells and in turn promote metastasis^14,15^. Neutrophilia is commonly accompanied by relative lymphocytopenia, representing a significant decline in the cell-mediated adaptive immune response. The NLR captures the balance between the detrimental effects of neutrophilia and the beneficial effects of lymphocyte-mediated adaptive immunity^16^.

Both systemic neutrophilia and lymphopenia are associated with reduced survival in cancer patients^17-20^. Many previous studies have examined the prognostic value of pre-treatment NLR^5,6^, and strong associations have consistently been demonstrated between high NLR and poor patient outcomes across many cancer types. Despite this, heterogeneity in the strength of association between NLR and overall survival has been observed^6^. Templeton et al. identified significant differences in pooled hazard ratios for association between high NLR and overall survival among 8 cancer types and between metastatic versus non-metastatic disease^6^, suggesting the prognostic potential of the NLR may be limited to specific patient subgroups.

Based on the rapid growth of existing literature in recent years, we conducted an updated meta-analysis including 228 studies (N=75,555 patients) relating NLR with overall survival in 18 cancer types. We quantify between-study heterogeneity and compare pooled hazard ratios by cancer type, as well as by region of study, therapy type, and cut-off for high NLR. We discuss publication bias and other potential sources of heterogeneity in the reported literature, and address the implications of this heterogeneity for translation of the NLR for clinical application.

## 2. Material and Methods

### 2.1. Data sources & extraction

The methods and results of the meta-analysis are reported in accordance with the PRISMA guidelines^21^. An electronic search of PubMed (including Medline 1974 to 1996) was conducted^1^ from inception to the end of 2017. Search terms included “neutrophil” “lymphocyte”, “ratio” and “NLR”, as well as “neutrophil:lymphocyte” and “neutrophil/lymphocyte”; details of the search strategy can be found in Supplementary Material S1a.

All titles identified by the search strategy were screened manually for association with cancerous diseases. After exclusion of non-cancer-associated manuscripts, remaining abstracts were screened for site-specific original research articles, which then proceeded to full text screening. The primary criteria for inclusion were: a) evaluation of the prognostic potential of the NLR as measured in peripheral blood, and b) availability of hazard ratios (HR) and 95% confidence intervals for overall survival based on multivariable regression analysis. Only studies in which NLR was obtained prior to the index treatment were included. We excluded studies that did not control for clinical and demographic covariates in the analysis (N=9), as well as studies published in non-English journals (N=28) and those in which NLR was reported as a continuous variable in multivariable analysis (N=10). All duplicates were omitted, including publications in which there was any potential overlap with patients in other included reports (Figure 1).

**Figure 1.**
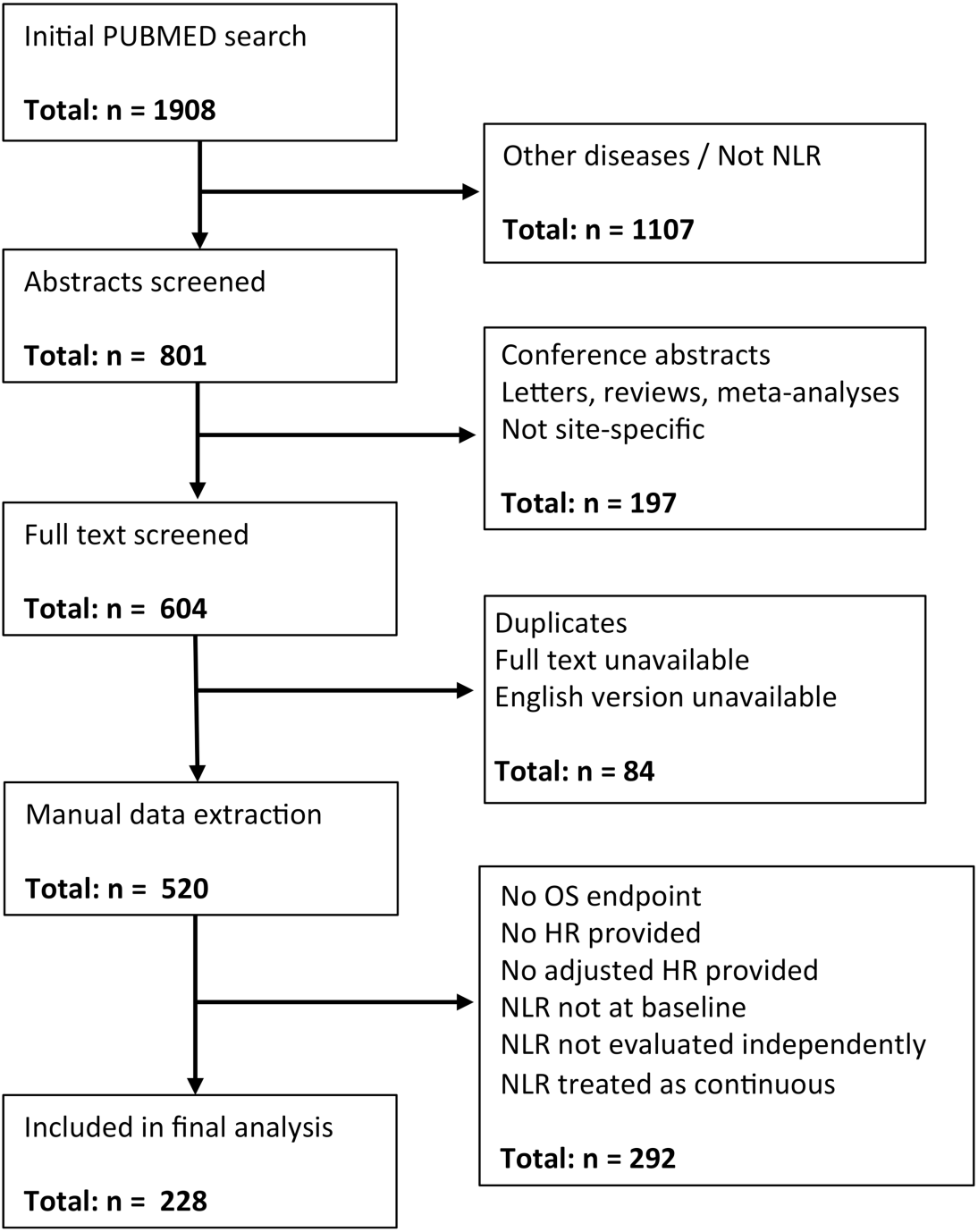
Flow chart of the study selection process. Figure 1 demonstrates the process of study selection that resulted in the final 228 studies for inclusion in the analysis. Exclusion criteria included: not associated with cancerous diseases; NLR as an acronym for something other than neutrophil-to-lymphocyte ratio; not a site-specific original research article; potential overlap of study subjects; no English full-text version available; no overall survival endpoint; no adjusted hazard ratio provided; NLR treated as continuous; NLR evaluated after initiation of treatment.

Overall survival (OS) was the primary outcome of interest. Disease-specific survival (DSS) was not assessed in the meta-analysis as only 6 cancer sites had multiple eligible manuscripts for analysis, as compared to 24 cancer sites with OS. Name of first author, year of publication, journal, region of study, number of patients included in analysis, age and sex distribution of patients, disease site, disease subtype(s), primary treatment type(s), average pre-treatment NLR, cut-off defining “high” NLR, hazard ratio for OS from multivariable analysis, and HR-associated 95% confidence intervals were extracted from each manuscript.

We identified cancer type of interest, chosen cut-off for high NLR, geographic region of study recruitment, and primary therapy type as study-level variables that were available for all contributing studies. Strata for cancer type (N=18) were defined for all cancers with 2 or more studies included in the meta-analysis. Cancers represented by only one study (N=6) were pooled into an “Other” category. NLR cut-points were <3.5 or ≥3.5, where the threshold of 3.5 was selected based on the average NLR cutoff across all studies included in the meta-analysis, weighted by number of participants per study (mean = 3.45, standard deviation = 0.85). Strata for region of study recruitment were Europe, North America, Asia, and Other. Categories for primary treatment included surgery, radiotherapy, chemotherapy and ‘other treatments’.

### 2.2. Statistical analysis

We quantified the range of HRs presented in all 228 studies and formally assessed between-study heterogeneity using Cochran’s Q test and Higgins I^2^ statistic. A p-value <0.05 in Cochran’s Q test and I^2^>50% were considered sufficient evidence of heterogeneity. Meta-regression was conducted to evaluate whether study-level variables, i.e. cancer type of interest, chosen cut-off for high NLR, geographic region of study recruitment, and primary therapy type, altered the magnitude of HRs.

We then assessed between-study heterogeneity within strata of each variable, for example, within all studies relating to lung cancer or within all studies in which patients received radiation therapy. If heterogeneity was observed, we used random effects models featuring restricted maximum-likelihood estimation to calculate the weighted and pooled HRs for this stratum by generic inverse variance. If no evidence of heterogeneity was found, fixed-effects (Mantel-Haenszel) models were used. The pooled HRs for each cancer type, region of study, therapy type, and for NLR cut-off above and below 3.5 were compared to identify subgroups in which NLR demonstrated stronger associations with survival (increased HR). Additionally, where heterogeneity in effect size was observed within a specific cancer type, we conducted univariate meta-regression analysis to evaluate the association between other study-level variables (region of study, therapy type, NLR cut-off) and study effect size within cancer type subgroups.

We evaluated publication bias by funnel plot visualization and the Egger bias test. This analysis was repeated for groups of studies stratified by region and by year of publication to evaluate whether certain groups were more prone to this form of bias.

All statistical tests were two-sided, and statistical significance was defined as p < 0.05. All statistical analyses were performed using SAS 9.4 (SAS Institute Inc., Cary, NC) and R version 3.3.2 (R core development team, Vienna, Austria).

## 3. Results

A total of 228 studies were included in the final meta-analysis, incorporating 75,555 individual patients. More than one study was available for 18 different cancers; 6 additional cancers had only one eligible study and thus were grouped together into the “Other” category. A summary of the included studies by cancer type is provided in **Table 1**, and a comprehensive list of all included studies can be found in **Supplementary Material S1b**. While individual-level data was not provided in any of the included manuscripts, where summary statistics were available for the distribution of age, sex or NLR within a study population, they have also been included in **Supplementary Material S1b**. These summary statistics varied dramatically across the included studies: median study-level patient ages ranged from 37 to 79, and median study-level patient NLR ranged from 1.7 to 7.3. Each study also defined a cut-off for high NLR, and these study-level cut-offs also varied significantly from 1.7 to 7.5. Weighted averages of these summary statistics by cancer type can be found in **Table 1**.

**Table 1.**
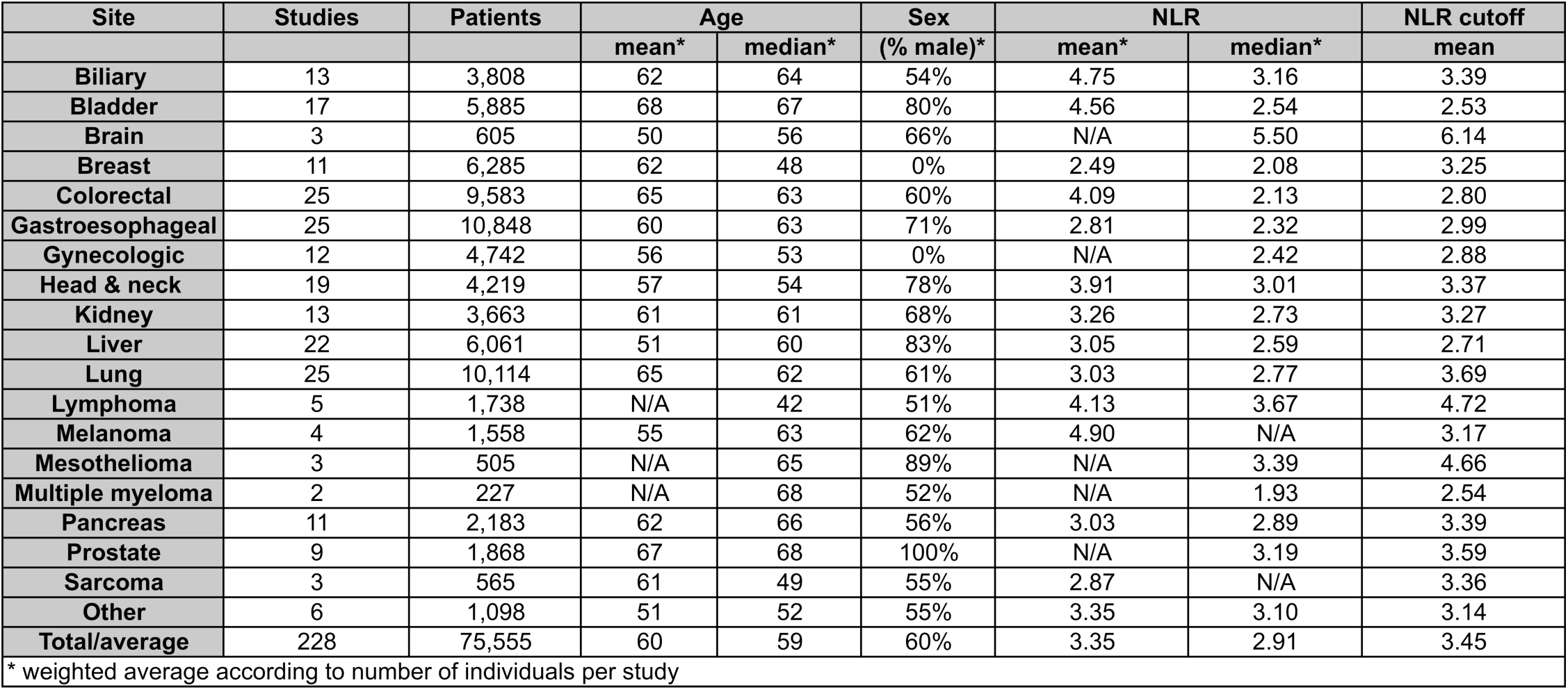
Summary of included studies. Table 1 presents a summary of studies (N=228) and patients (N=75,555) included in the meta-analysis, grouped by cancer type. Summary statistics represent a weighted average value based on the number of participants in each contributing study.

| Site | Studies | Patients | Age |  | Sex<br>(% male)* | NLR |  | NLR cutoff<br>mean |
| --- | --- | --- | --- | --- | --- | --- | --- | --- |
|  |  |  | mean* | median* |  | mean* | median* |  |
| Biliary | 13 | 3,808 | 62 | 64 | 54% | 4.75 | 3.16 | 3.39 |
| Bladder | 17 | 5,885 | 68 | 67 | 80% | 4.56 | 2.54 | 2.53 |
| Brain | 3 | 605 | 50 | 56 | 66% | N/A | 5.50 | 6.14 |
| Breast | 11 | 6,285 | 62 | 48 | 0% | 2.49 | 2.08 | 3.25 |
| Colorectal | 25 | 9,583 | 65 | 63 | 60% | 4.09 | 2.13 | 2.80 |
| Gastroesophageal | 25 | 10,848 | 60 | 63 | 71% | 2.81 | 2.32 | 2.99 |
| Gynecologic | 12 | 4,742 | 56 | 53 | 0% | N/A | 2.42 | 2.88 |
| Head & neck | 19 | 4,219 | 57 | 54 | 78% | 3.91 | 3.01 | 3.37 |
| Kidney | 13 | 3,663 | 61 | 61 | 68% | 3.26 | 2.73 | 3.27 |
| Liver | 22 | 6,061 | 51 | 60 | 83% | 3.05 | 2.59 | 2.71 |
| Lung | 25 | 10,114 | 65 | 62 | 61% | 3.03 | 2.77 | 3.69 |
| Lymphoma | 5 | 1,738 | N/A | 42 | 51% | 4.13 | 3.67 | 4.72 |
| Melanoma | 4 | 1,558 | 55 | 63 | 62% | 4.90 | N/A | 3.17 |
| Mesothelioma | 3 | 505 | N/A | 65 | 89% | N/A | 3.39 | 4.66 |
| Multiple myeloma | 2 | 227 | N/A | 68 | 52% | N/A | 1.93 | 2.54 |
| Pancreas | 11 | 2,183 | 62 | 66 | 56% | 3.03 | 2.89 | 3.39 |
| Prostate | 9 | 1,868 | 67 | 68 | 100% | N/A | 3.19 | 3.59 |
| Sarcoma | 3 | 565 | 61 | 49 | 55% | 2.87 | N/A | 3.36 |
| Other | 6 | 1,098 | 51 | 52 | 55% | 3.35 | 3.10 | 3.14 |
| Total/average | 228 | 75,555 | 60 | 59 | 60% | 3.35 | 2.91 | 3.45 |

\* weighted average according to number of individuals per study

The association between NLR and OS was significant in 195 studies (86%). The HRs for baseline NLR and overall survival across all included studies ranged from 1.02 to above 8, with an overall pooled HR of 1.75 [1.69-1.82]. Cochran’s Q test also indicated a high level of heterogeneity across studies (Q = 3696, p<0.0001), as did Higgins I^2^ statistic (I^2^ = 94.5%).

Forest plots of pooled HRs for strata of cancer type, region of study, therapy type, and NLR cutoff can be found in **Figure 2**. There was considerable evidence of heterogeneity across each of these four variables, with pCochran < 0.001 and I^2^ > 80% in all cases. For cancer type, the pooled HR for the association between baseline NLR and OS was weakest in patients with brain cancer (HR = 1.05, 95% CI = 1.01-1.1) and strongest in patients with melanoma (HR = 2.65, 95% CI = 1.28-5.51). European studies (HR = 1.84, 95% CI = 1.80-1.88), studies in which radiation was the primary treatment modality (HR = 1.96, 95% CI = 1.84-2.08), and studies in which the threshold for high NLR was above the overall median of 3.5 (HR = 1.91, 95% CI = 1.87-1.95) all exhibited a higher pooled HR than other strata within their respective variables. Despite this, with the exception of brain cancer (p=0.04), meta-regression analyses for the association between study effect size and cancer type, region of study, therapy type and NLR cutoff resulted in non-significant p-values for all strata (not shown).

**Figure 2.**
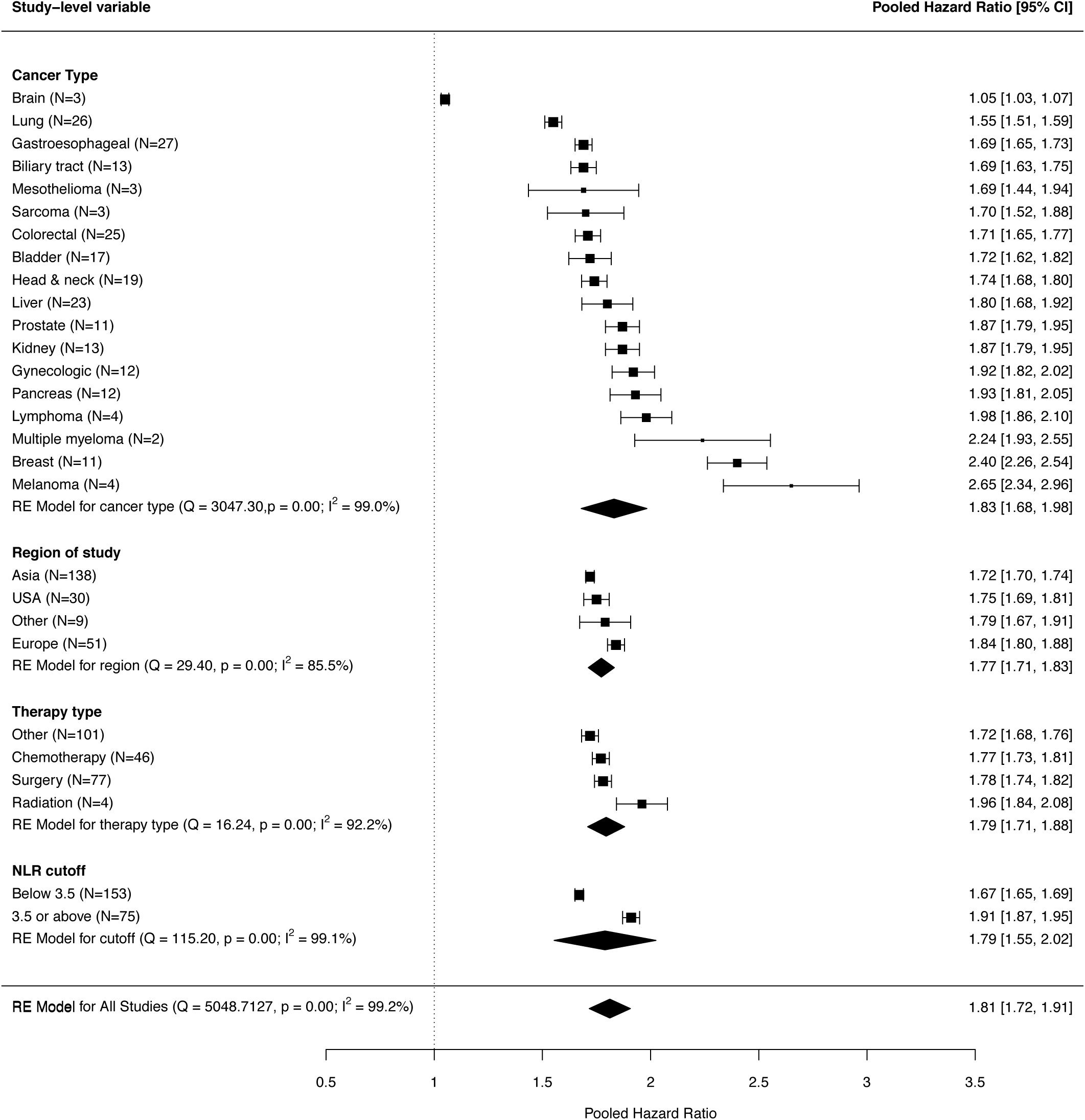
Pooled hazard ratios. Figure 2 demonstrates pooled hazard ratios for each stratum of four variables of interest: cancer type, region of study, therapy type and NLR cut-off. N represents the number of studies included in the calculation of the pooled point estimate. The size of the point estimate marker for each stratum is proportional to the number of patients included. Strata are ranked by magnitude of hazard ratio.

Forest plots for all studies sorted by cancer type, and the models used for calculating these weighted and pooled hazard ratios can be found in **Supplementary Material S1c**. Heterogeneity of effect was observed within cancer types, including studies of bladder, breast, gynecologic, kidney, liver, lung and pancreatic cancer, as well as melanoma and mesothelioma. Significant findings from univariate meta-regression analysis within these cancer type subgroups are summarized in **Figure 3**. The choice of cut-off for high NLR appeared to moderate effect size in studies of breast cancer, liver cancer and melanoma. Therapy type was also significantly associated with effect size in pancreatic cancer, and region of study was significantly associated with effect size in melanoma. All other meta-regression associations were not significant (not shown).

**Figure 3.**
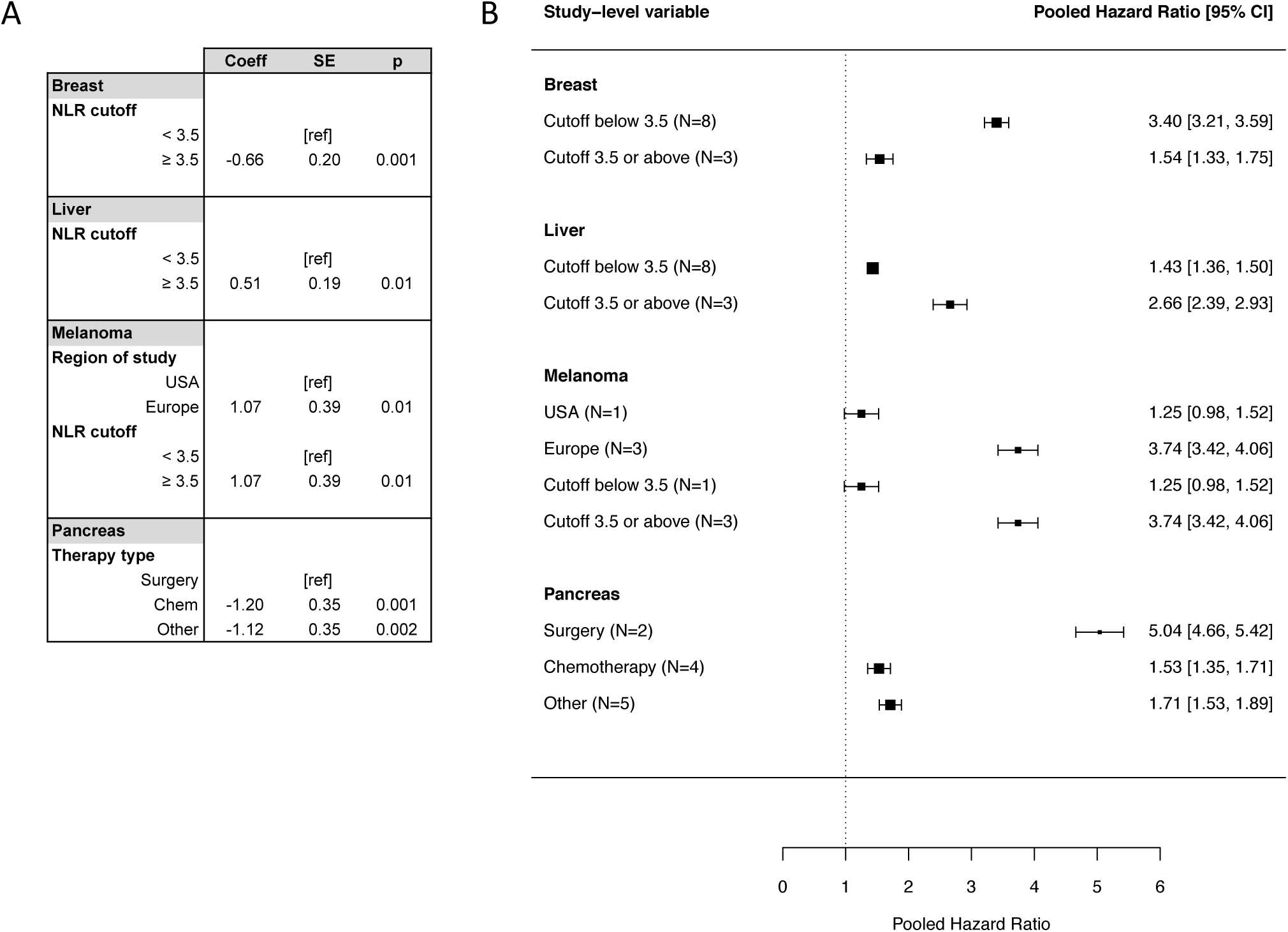
Meta-regression results. Panel A of Figure 3 demonstrates meta-regression coefficients and p-values for all significant associations between study effect size and study-level variables (region of study, therapy type, NLR cutoff) within cancer type subgroups. Panel B shows an alternative representation of results shown in A (significant differences in study effect size according to study-level variables within cancer type subgroups) via pooled HRs. Note that the one study from the USA is also the only study with an NLR cut-off below 3.5, leading to identical meta-regression coefficients and pooled HRs.

The funnel plot (**Figure 4**) and Egger regression test (z=14.77, p<0.0001) suggested the presence of publication bias. Publication bias was also significant (p<0.0001) after stratifying by year of publication (strata defined as 2013 and earlier, 2014, 2015, 2016 and 2017) and region of study, with the exception of the “other” region category - only 9 studies were available for analysis within this stratum (not shown).

**Figure 4.**
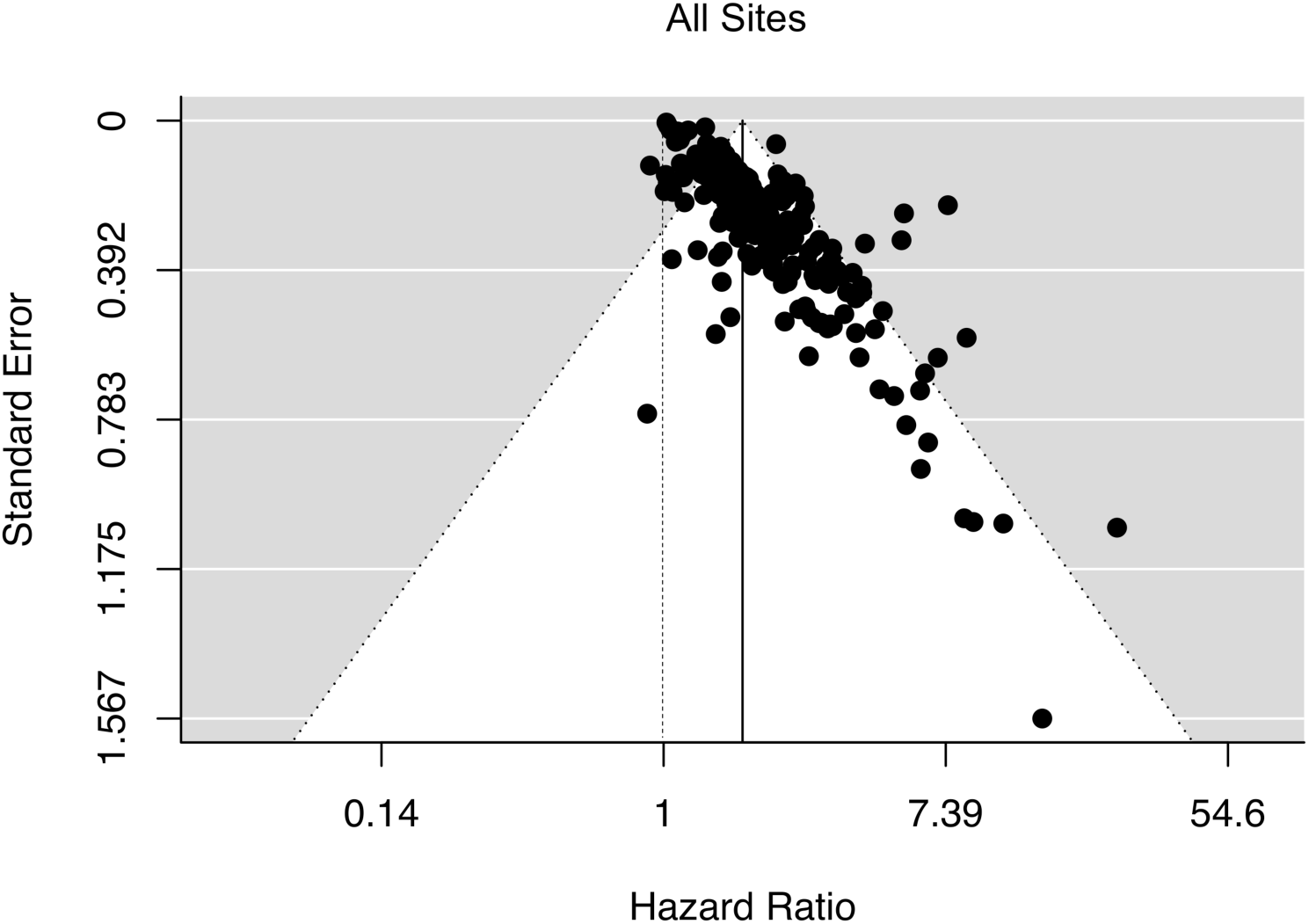
Funnel plot. Figure 4 presents a funnel plot to assess publication bias across studies included in meta-analysis. HR of high NLR and overall survival is presented on the horizontal axis and standard error for this HR is presented on the vertical axis. Each study is represented by a single point; the solid vertical line represents the pooled effect estimate for all studies.

## 4. Discussion

As previously noted by Templeton et al.^6^, the strength of association between NLR and overall survival varies significantly between published studies. In our extended meta-analysis, the pooled hazard ratio for the association between NLR and OS was highest in studies of melanoma and lowest in studies of brain cancer. While this might initially suggest an association between the immunogenicity of the cancer and prognostic potential of the NLR, breast cancer also demonstrates a high pooled HR, and in lung cancer this association is comparably weak. Radiation as primary treatment also demonstrated a large pooled effect size as compared to that for all studies; other treatment types and region of study demonstrated only negligible differences.

While clear differences in strength of association are evident between study subgroups, a high level of inter-study heterogeneity in effect size is also observed *within* many study subgroups, leading to large confidence intervals for pooled estimates. Some of this variation may be attributable to clinical differences that were captured in insufficient detail by the meta-analysis, for example variation in histologic disease subtypes within one broad cancer type, multi-modality treatment regimens, or within-region variation in race and ethnicity. The tendency of authors to use minimum p-value methods for identifying optimal cutoffs for high NLR could also contribute to this observed heterogeneity; our results suggested that a high threshold for classifying high-risk NLR led to a significantly higher pooled HR. Further inflation of summary point estimates could be resulting from publication bias, specifically the underreporting of studies with small effect sizes. As such, the findings from this and other meta-analyses of the NLR in cancer patients should be interpreted with caution.

While a large body of literature exists on the prognostic potential of the NLR, the translational value of this marker in the clinical setting remains to be determined. Based on the results of the present study, the NLR has greater prognostic value in certain cancer types and for different therapeutic regimens. Some observations of the earlier meta-analysis by Templeton et al. are supported in the present study, including that biliary tract, gastroesophageal, and lung cancers exhibit relatively weaker associations between NLR and OS than kidney and pancreatic cancers. However, some adjustments to the ranking of HR by cancer type are evident in this expanded analysis: liver cancer no longer has a low NLR relative to other sites, for example. These discrepancies could be attributable to the increased number of studies, or could be a result of the inherent weaknesses outlined above. In either case, efforts are needed to comprehensively examine population subgroups in which NLR has maximum prognostic power, and to identify clinically meaningful thresholds for risk stratification within these populations. With this knowledge, prospective evaluation of the prognostic power of the NLR within these groups can be conducted to determine whether clinical implementation of the NLR as a prognostic tool is a realistic and attainable goal.

## Data Availability

The manuscript is a meta-analysis of published work, no new data were collected or analyzed in this study.

## Footnotes

1 Last searched: July 2018

## Notes

### Competing Interest Statement

The authors have declared no competing interest.

### Funding Statement

Dr. Rachel Howard was supported by a Ruth L. Kirschstein Institutional National Research Service award from the National Institutes of Health (T32 CA147832) during the preparation of this manuscript. The National Institutes of Health had no direct role in the design of this study, nor the execution, analyses interpretation of the data or decision to submit results. This research did not receive any specific grant from funding agencies in the public, commercial, or not-for-profit sectors.

